# Predicting loss of independence and mortality in frontotemporal lobar degeneration syndromes

**DOI:** 10.1101/2020.02.11.20022061

**Authors:** Alexander G. Murley, Matthew A. Rouse, Ian Coyle-Gilchrist, P Simon Jones, Win Li, Julie Wiggins, Claire Lansdall, Patricia Vazquez Rodriguez, Alicia Wilcox, Karalyn Patterson, James B. Rowe

## Abstract

**Objective:** To test the hypothesis that in syndromes associated with frontotemporal lobar degeneration, behavioural impairment predicts loss of functional independence and motor clinical features predict mortality, irrespective of syndrome subtype.

**Method:** We used a transdiagnostic approach to survival in an epidemiological cohort, testing the association between clinical features, independence and survival in patients with clinical diagnoses of behavioural variant frontotemporal dementia (n=64), non-fluent variant primary progressive aphasia (n=36), semantic variant primary progressive aphasia (n=25), progressive supranuclear palsy (n=101) and corticobasal syndrome (n=68). A principal components analysis identified six dimensions of clinical features. Using Cox proportional hazards and logistic regression we identified the association between each of these dimensions and functionally independent survival (time from clinical assessment to care home admission), and absolute survival (time to death). Analyses adjusted for the covariates of age, gender and diagnostic group. Secondary analysis excluded specific diagnostic groups.

**Results:** Behavioural disturbance, including impulsivity and apathy, was associated with reduced functionally independent survival (OR 2.46, p<0.001), even if patients with bvFTD were removed from the analysis. Motor impairments were associated with reduced absolute survival, even if patients with PSP and CBS were removed from the analysis.

**Conclusion:** Our results may help individualised prognostication and planning of disease-modifying trials and support a transdiagnostic approach to symptomatic treatment trials in patients with clinical syndromes associated with frontotemporal lobar degeneration.

## Introduction

Prognosis in syndromes associated with frontotemporal lobar degeneration (FTLD) is highly variable and difficult to predict. Disease duration is not fully explained by the diagnostic categorisation to behavioural variant frontotemporal dementia (bvFTD), non-fluent (nfvPPA) or semantic (svPPA) variants of primary progressive aphasia, progressive supranuclear palsy (PSP) or corticobasal syndrome (CBS) [1–5]. Better prognostic models would aid both trial design and clinical management.

The syndromes caused by frontotemporal lobar degeneration (FTLD) have highly heterogenous and overlapping clinical features. Our hypothesis was that a subset of clinical features, represented across the spectrum of disorders might explain variation in functional independence and survival [6,7]. We therefore adopted a transdiagnostic approach, increasingly used in psychiatric and neurological diseases, to identify prognostic clinical features across the FTLD syndrome spectrum [8,9]. Previous work has identified that features of motor neurone disease reduce the prognosis in bvFTD [1,3,10], while dysphagia and cognitive impairment worsen prognosis in PSP-Richardson’s syndrome [11]. Here we focus on cognitive, behavioural and motor features of disease.

Mortality is a definite endpoint in FTD, PSP and CBS. However, these disorders also engender dependency and caregiver burden [5,12–15]. Community-based studies suggest increased dependency, due to cognitive or physical impairment, predicts care home admission [16,17]. We use care home admission as a disease endpoint that indirectly represents loss of functional independence [5,14].

We test the hypothesis that clinical features predict prognosis over and above the diagnostic label. Specifically, we test whether behavioural impairments increase the risk of care home admission while motor impairments are associated with earlier mortality.

## Methods

### Participant recruitment and clinical review

Survival data were collected for all participants in the PIPPIN study (Pick’s disease and Progressive Supranuclear Palsy Prevalence and Incidence), a cross-sectional epidemiological study, details of which have been previously reported [2,7]. This study enrolled via multisource referral, all patients with a designated syndrome associated with frontotemporal lobar degeneration syndrome living in the UK counties of Cambridgeshire and Norfolk over two 24-month periods (2013-14, 2017-18).

We use current consensus terminology: frontotemporal lobar degeneration refers to neuropathological classification. FTLD is associated with a range of clinical syndromes that include behavioural variant frontotemporal dementia (bvFTD), non-fluent (nfvPPA) and semantic (svPPA) variants of primary progressive aphasia, progressive supranuclear palsy (PSP) and corticobasal syndrome (CBS). We grouped together patients with logopenic variant primary progressive aphasia and primary progressive aphasia who did not meet criteria for one of the three PPA subtypes, noting that both these groups commonly have underlying Alzheimer’s disease [18]. We assessed in person 310 of the 365 patients identified as alive and in area in the time windows. A clinical, cognitive and language assessment recorded the presence or absence of clinical symptoms and signs included in the current diagnostic criteria for FTLD syndromes [19–23], plus cognitive assessment using the Addenbrookes Cognitive Examination – Revised (ACER) [24] and carer interview using the Cambridge Behavioural Inventory – Revised (CBIR) [25].

We recorded dates of care home admission and death from each participant’s NHS Summary Care Record. This database includes information on the address and date of death of every UK resident, minimising loss to follow up. We defined a care home as an institution registered with the UK government to provide residential and/or nursing care. All participants provided written informed consent or, if they lacked capacity to consent then their next of kin was consulted using the ‘personal consultee’ process according to UK law. The study had ethical approval from the Cambridge Central Research Ethics Committee (REC 12/EE/0475).

### Statistical analysis

We employed a transdiagnostic, data-driven approach using principal component analysis to identify syndrome dimensions of covarying clinical features. Forty-five clinical features were combined into twenty-four groups of related features by summing the number of present features in each group (the clinical features and groups are given in Supplementary Materials). The clinical feature group scores, ACER and CBIR results were standardised into z scores then entered into a principal component analysis. We identified six components using Cattell’s criterion and then performed varimax rotation.

We used a Cox proportional hazards regression analysis to test the association between these six clinical syndrome components and the time from clinical assessment to death (covariates of age, gender and disease group). This allows all participants to be included in the survival analysis, censoring participants who failed to reach the end point (death). The predictor variables (subject-weightings on each syndrome dimension) were z scored to aid interpretation. If a syndrome dimension closely resembled typical features of a specific diagnostic group we repeated the Cox proportional hazards regression analysis without that group.

Next, we tested the association between the syndrome dimensions and time to care home admission using logistic regression, with the binary outcome of care home admission by 2 years from study assessment. Patients in a care home at study assessment or those with incomplete follow up were excluded from this analysis. We used logistic rather than Cox proportional hazards regression for two reasons: first, to allow assessment of care home admission risk independent of mortality and second, because it could be argued that the risk of care admission does not remain constant over time (an assumption of Cox hazards regression). All analyses were performed in Matlab 2018b (Mathworks, USA). Kaplan-Meier curves were plotted using the *MatSurv* function (https://github.com/aebergl/MatSurv).

## Results

365 patients with a designated diagnosis were identified as alive in region within the time windows, of whom 310 were assessed in person by the study team (bvFTD n=64, nfvPPA n=36, svPPA n=25, other PPA n=16, PSP n=101, CBS n=68). The epidemiological, phenotypic, neuropsychological and imaging characteristics of this cohort at baseline have been published previously [2,7,26] Summary demographic and survival results are shown in Table 1. At the censor date (1 August 2019) 169 FTLD patients (54.5%) had been admitted to a care home and 200 patients (64.5%) had died. Most patients were admitted to a care home before they died (131/200, 65.5%).

**Table 1:**
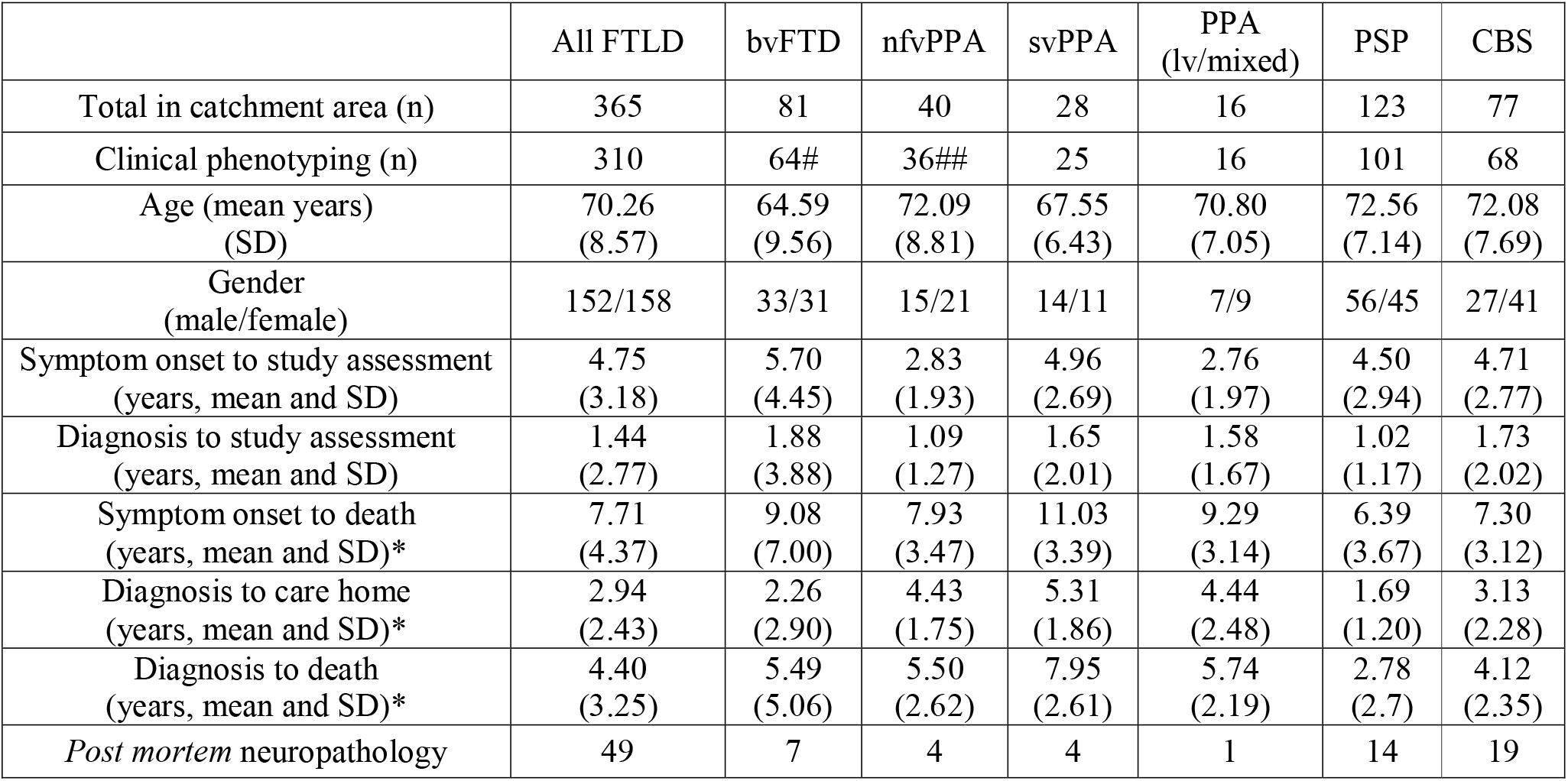
Demographics of the study cohort. *Subgroup of cohort with complete follow up. 6 patients were living in a care home at diagnosis. #12 patients with bvFTD had motor neuron disease. ##1 patient with nfvPPA had motor neurone disease.

|  | All FTLD | bvFTD | nfvPPA | svPPA | PPA<br>(lv/mixed) | PSP | CBS |
| --- | --- | --- | --- | --- | --- | --- | --- |
| Total in catchment area (n) | 365 | 81 | 40 | 28 | 16 | 123 | 77 |
| Clinical phenotyping (n) | 310 | 64# | 36## | 25 | 16 | 101 | 68 |
| Age (mean years)<br>(SD) | 70.26<br>(8.57) | 64.59<br>(9.56) | 72.09<br>(8.81) | 67.55<br>(6.43) | 70.80<br>(7.05) | 72.56<br>(7.14) | 72.08<br>(7.69) |
| Gender<br>(male/female) | 152/158 | 33/31 | 15/21 | 14/11 | 7/9 | 56/45 | 27/41 |
| Symptom onset to study assessment<br>(years, mean and SD) | 4.75<br>(3.18) | 5.70<br>(4.45) | 2.83<br>(1.93) | 4.96<br>(2.69) | 2.76<br>(1.97) | 4.50<br>(2.94) | 4.71<br>(2.77) |
| Diagnosis to study assessment<br>(years, mean and SD) | 1.44<br>(2.77) | 1.88<br>(3.88) | 1.09<br>(1.27) | 1.65<br>(2.01) | 1.58<br>(1.67) | 1.02<br>(1.17) | 1.73<br>(2.02) |
| Symptom onset to death<br>(years, mean and SD)* | 7.71<br>(4.37) | 9.08<br>(7.00) | 7.93<br>(3.47) | 11.03<br>(3.39) | 9.29<br>(3.14) | 6.39<br>(3.67) | 7.30<br>(3.12) |
| Diagnosis to care home<br>(years, mean and SD)* | 2.94<br>(2.43) | 2.26<br>(2.90) | 4.43<br>(1.75) | 5.31<br>(1.86) | 4.44<br>(2.48) | 1.69<br>(1.20) | 3.13<br>(2.28) |
| Diagnosis to death<br>(years, mean and SD)* | 4.40<br>(3.25) | 5.49<br>(5.06) | 5.50<br>(2.62) | 7.95<br>(2.61) | 5.74<br>(2.19) | 2.78<br>(2.7) | 4.12<br>(2.35) |
| Post mortem neuropathology | 49 | 7 | 4 | 4 | 1 | 14 | 19 |

There was high variability in the time from diagnosis to care home admission or death in all groups (Figure 1). Life expectancy differed between groups (ANOVA, F1,5 =10.41, p<0.001). This was primarily due to longer life expectancy in svPPA compared to PSP (mean difference 5.24 years, p<0.001), CBS (3.83 years, p<0.001) and bvFTD (2.69 years, p=0.047). PSP patients also had a worse prognosis compared to bvFTD (mean difference 2.55 years, p<0.001) and nfvPPA (2.54 years, p<0.001). Thirteen patients with FTD-MND had a lower mean time between diagnosis and death than the whole bvFTD cohort (2.67 vs 5.49 years). Post hoc tests were Bonferroni corrected, and a table of all FTLD subgroup comparisons is in supplementary materials.

**Figure 1.**
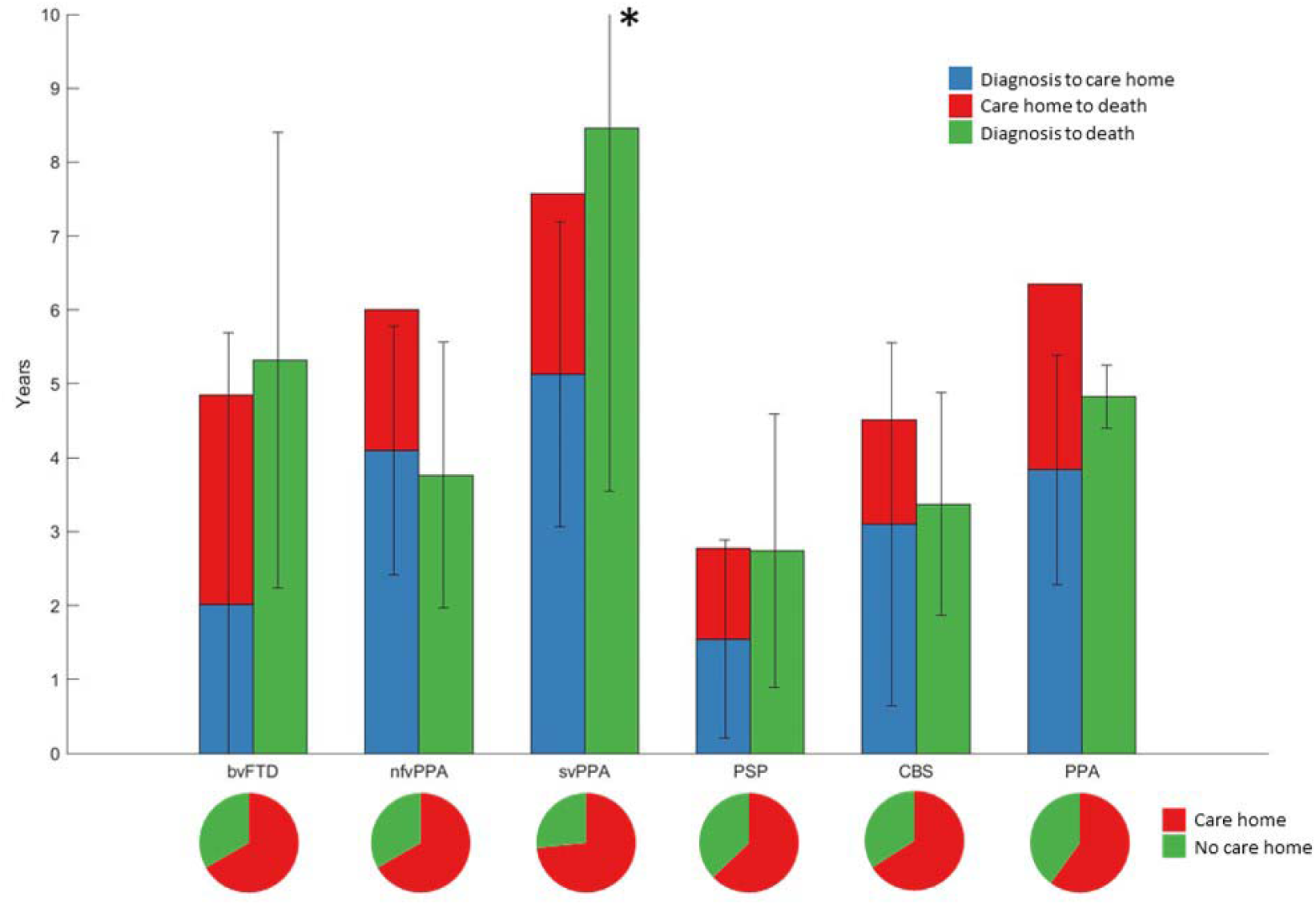
Survival in frontotemporal lobar degeneration syndromes. The barplot shows disease duration in FTLD syndromes in patients with complete follow up from disease onset to death. Survival in each FTLD subgroup is shown grouped by care home vs no care home admission. The pie charts show proportion of each FTLD subgroup admitted to a care home during the disease course.

Using principal component analysis, we identified six clinical symptom dimensions (Table 2). An individual’s score on each dimension showed the extent to which they expressed that clinical phenotype. Syndrome dimension 1 reflected high clinician and carer rating of behavioural impairment. Syndrome dimension 2 reflected cognitive impairment, with contribution from all ACER subscales and carer ratings of memory and everyday skills. Syndrome dimension 3 mirrored a PSP-RS-like motor phenotype, with positive loadings reflecting symmetrical parkinsonism, falls and supranuclear gaze palsy. Negative loadings on this dimension reflected semantic language impairment. The fourth syndrome dimension represented asymmetrical parkinsonism, myoclonus and dystonia with cortical features of alien limb syndrome, apraxia and cortical sensory loss. Syndrome dimension five was driven by language impairments including speech apraxia, loss of syntactic comprehension and impaired repetition. Syndrome dimension six reflected carer ratings of low mood and abnormal beliefs. There was a spread of scores across FTLD subgroups onto these symptom domains.

**Table 2.** (previous page): - Varimax-rotated component matrix from principal component analysis. ACER: Addenbrooke’s Cognitive Examination – Revised. CBI: Cambridge Behavioural Inventory. Positive loadings indicate worse performance or presence of symptoms, except for ACER where negative loadings indicate worse performance. Factor loadings above 0.4 or below −0.4 shown in bold.

|  | Syndrome<br>Dimension<br>1 | Syndrome<br>Dimension<br>2 | Syndrome<br>Dimension<br>3 | Syndrome<br>Dimension<br>4 | Syndrome<br>Dimension<br>5 | Syndrome<br>Dimension<br>6 |
| --- | --- | --- | --- | --- | --- | --- |
| Disinhibition | **0.7399** | 0.0774 | -0.0790 | -0.1423 | -0.1576 | 0.1102 |
| Apathy | **0.5486** | 0.0763 | **0.4276** | 0.1221 | -0.1919 | 0.1450 |
| Loss of sympathy or empathy | **0.7022** | 0.1278 | 0.0044 | -0.0981 | -0.1278 | 0.0205 |
| Stereotyped/compulsive<br>behaviour | **0.5789** | 0.2890 | -0.3103 | -0.1798 | -0.0536 | 0.2444 |
| Hyperorality/dietary change | **0.6234** | 0.0459 | -0.2198 | -0.1270 | -0.0932 | 0.0705 |
| Executive dysfunction | **0.5458** | 0.1440 | 0.1176 | -0.0176 | -0.0476 | 0.3155 |
| CBI - Abnormal behaviour | **0.7497** | 0.1251 | -0.0348 | -0.0545 | -0.0898 | -0.3164 |
| CBI - Mood | **0.5485** | 0.1013 | 0.0039 | 0.1709 | -0.0077 | **-0.5067** |
| CBI - Eating habits | **0.7647** | 0.0394 | -0.0486 | -0.0500 | -0.0074 | -0.2188 |
| CBI - Sleep | **0.4569** | -0.0259 | 0.2813 | 0.1776 | -0.0488 | **-0.4043** |
| CBI - Motor behaviour | **0.7056** | -0.0161 | -0.1997 | -0.1174 | 0.0726 | -0.2513 |
| CBI - Motivation | **0.7075** | 0.2981 | 0.1511 | 0.0500 | -0.1157 | -0.1488 |
| ACER - Attention/orientation | -0.1510 | **-0.9002** | 0.0922 | 0.0463 | -0.0248 | 0.0724 |
| ACER - Memory | -0.1124 | **-0.8258** | 0.3410 | 0.1746 | -0.0440 | 0.0770 |
| ACER - Fluency | -0.1784 | **-0.7576** | 0.1183 | 0.1772 | -0.0744 | -0.1227 |
| ACER - Language | -0.0805 | **-0.8460** | 0.3405 | 0.1337 | 0.0314 | 0.0238 |
| ACER - Visuospatial | -0.0532 | **-0.8299** | -0.1642 | -0.1818 | -0.0460 | 0.1518 |
| CBI - Memory | **0.4864** | **0.5544** | -0.2152 | -0.0392 | 0.0199 | -0.3158 |
| CBI - Everyday skills | **0.4086** | **0.5214** | 0.3309 | 0.3198 | 0.0727 | -0.1291 |
| Symmetrical parkinsonism | 0.0127 | -0.0415 | **0.7676** | -0.3673 | 0.0655 | -0.0362 |
| Axial rigidity | -0.0156 | -0.1158 | **0.8077** | 0.0425 | -0.0231 | 0.0669 |
| Poor levodopa responsiveness | -0.1307 | -0.1057 | **0.6757** | 0.0873 | -0.0629 | -0.0536 |
| Postural instability | -0.0504 | -0.1069 | **0.7719** | 0.1690 | -0.0640 | -0.0241 |
| Supranuclear gaze palsy | -0.0938 | -0.1144 | **0.8132** | 0.1045 | -0.0473 | 0.0526 |
| CBI - Self care | 0.3459 | 0.3996 | **0.4524** | 0.3626 | -0.0628 | -0.0775 |
| Impaired semantics | 0.1510 | 0.3067 | **-0.5187** | -0.2377 | 0.0353 | 0.1970 |
| Asymmetrical parkinsonism | -0.0652 | -0.0843 | 0.0700 | **0.8343** | -0.0627 | 0.0202 |
| Asymmetrical dystonia | 0.0282 | -0.0673 | 0.0899 | **0.8300** | -0.0550 | 0.1061 |
| Asymmetrical myoclonus | -0.0340 | -0.0148 | -0.0493 | **0.6830** | 0.1012 | 0.0621 |
| Limb apraxia | -0.1292 | -0.0590 | 0.1252 | **0.5274** | **0.5056** | -0.0233 |
| Cortical sensory loss | -0.1462 | -0.0201 | 0.0584 | **0.5635** | 0.2569 | -0.2505 |
| Alien limb syndrome | -0.0363 | -0.0066 | 0.0504 | **0.5423** | 0.0749 | -0.1367 |
| Symmetrical myoclonus | -0.0658 | -0.0350 | 0.0044 | -0.0093 | **0.5132** | -0.3228 |
| Agrammatic, apraxic speech | -0.1224 | 0.1231 | -0.0369 | 0.1137 | **0.7667** | 0.2703 |
| Logopenic speech | -0.0659 | 0.0268 | -0.1180 | -0.0461 | **0.7752** | 0.0415 |
| CBI - Beliefs | 0.1830 | 0.2358 | 0.0220 | -0.0019 | 0.0093 | **-0.5919** |
| Symmetrical dystonia | 0.1134 | 0.1176 | 0.3325 | -0.1741 | 0.2010 | 0.0008 |
| Orobuccal apraxia | -0.1225 | -0.0170 | -0.0012 | 0.2822 | 0.3727 | -0.0267 |
| Visuospatial deficits | -0.1863 | 0.1862 | -0.0106 | 0.2386 | 0.3650 | -0.2559 |
| Motor neuron disease | 0.2615 | -0.1304 | -0.1584 | -0.0617 | -0.1683 | 0.0556 |

Cox proportional hazards regression indicated that syndrome dimensions 3 and 4 and age at clinical assessment were associated with shorter time to death (Table 3). Syndrome dimension 3 remained a significant predictor of death after PSP was removed (HR 2.30, CI 1.50-3.52, p<0.001). Absolute survival (time from assessment to death) differed between participants in high, medium and low severity tertiles for syndrome dimensions 3 (Figure 2A) and 4 (Figure 2B) severity score. This effect persisted after removing the highest scoring FTLD subgroups, PSP for syndrome dimension 3 (logrank p<0.001) and CBS for syndrome dimension 4 (logrank p<0.001).

**Table 3:** Cox proportional hazards model of time from study assessment to death.

|  | Hazard ratio | Hazard ratio (CI) | Coefficient | SE | P value |
| --- | --- | --- | --- | --- | --- |
| **Age** | **1.04** | **(1.02-1.06)** | **0.04** | **0.01** | **<0.01** |
| Gender | 1.18 | (0.87-1.59) | 0.16 | 0.16 | 0.29 |
| Diagnosis 1 | 0.64 | (0.23-1.80) | -0.45 | 0.53 | 0.40 |
| Diagnosis 2 | 0.87 | (0.48-1.57) | -0.14 | 0.3 | 0.64 |
| Diagnosis 3 | 1.16 | (0.53-2.54) | 0.15 | 0.4 | 0.71 |
| Diagnosis 4 | 0.99 | (0.5-1.97) | -0.01 | 0.35 | 0.97 |
| Diagnosis 5 | 0.65 | (0.25-1.64) | -0.44 | 0.48 | 0.36 |
| Syndrome dimension 1 | 1.23 | (0.98-1.55) | 0.21 | 0.12 | 0.07 |
| Syndrome dimension 2 | 1.15 | (0.99-1.35) | 0.14 | 0.08 | 0.07 |
| **Syndrome dimension 3** | **1.97** | **(1.41-2.75)** | **0.68** | **0.17** | **<0.01** |
| **Syndrome dimension 4** | **1.31** | **(1.07-1.61)** | **0.27** | **0.1** | **<0.01** |
| Syndrome dimension 5 | 0.87 | (0.73-1.03) | -0.14 | 0.09 | 0.12 |
| Syndrome dimension 6 | 0.88 | (0.75-1.03) | -0.13 | 0.08 | 0.12 |

**Figure 2.**
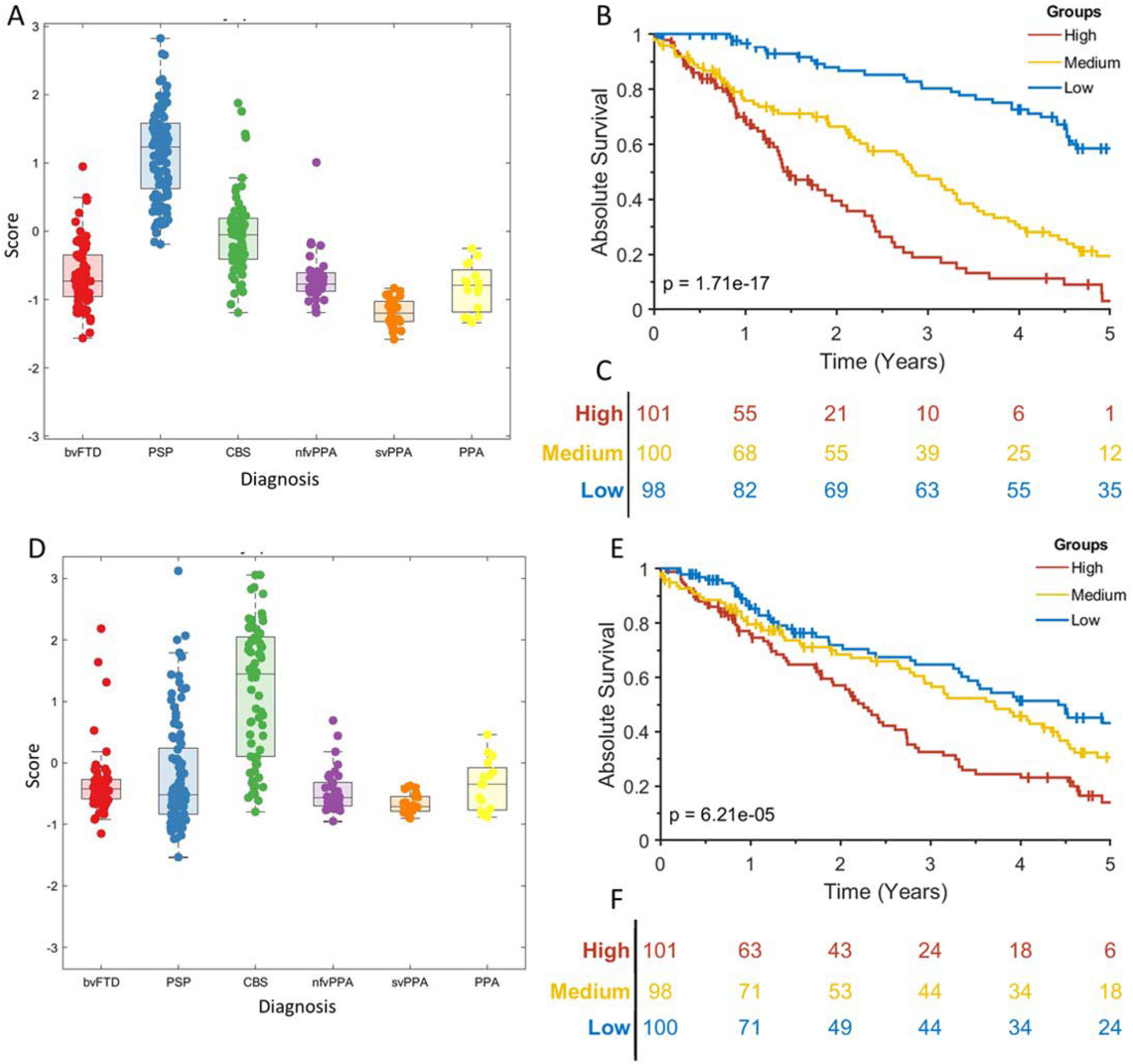
Absolute survival (time to death) in frontotemporal lobar degeneration syndrome. 2A: Scatterboxplot of individual’s scores on syndrome dimension three, grouped by FTLD syndrome subtype. 2B: Kaplan Meier survival curve for high, medium and low scoring tertiles for syndrome dimension three. The p value is from a log rank test of the null hypothesis of no difference in survival between all groups. Vertical lines show censored data. 2C: At risk table for the data shown in 2B. 2D: Scatterboxplot of individual’s scores on syndrome dimension four 2E: Kaplan Meier survival curve for high, medium and low scoring tertiles for syndrome dimension four. 2F: At risk table for the data shown in 2E.

**Figure 3.**
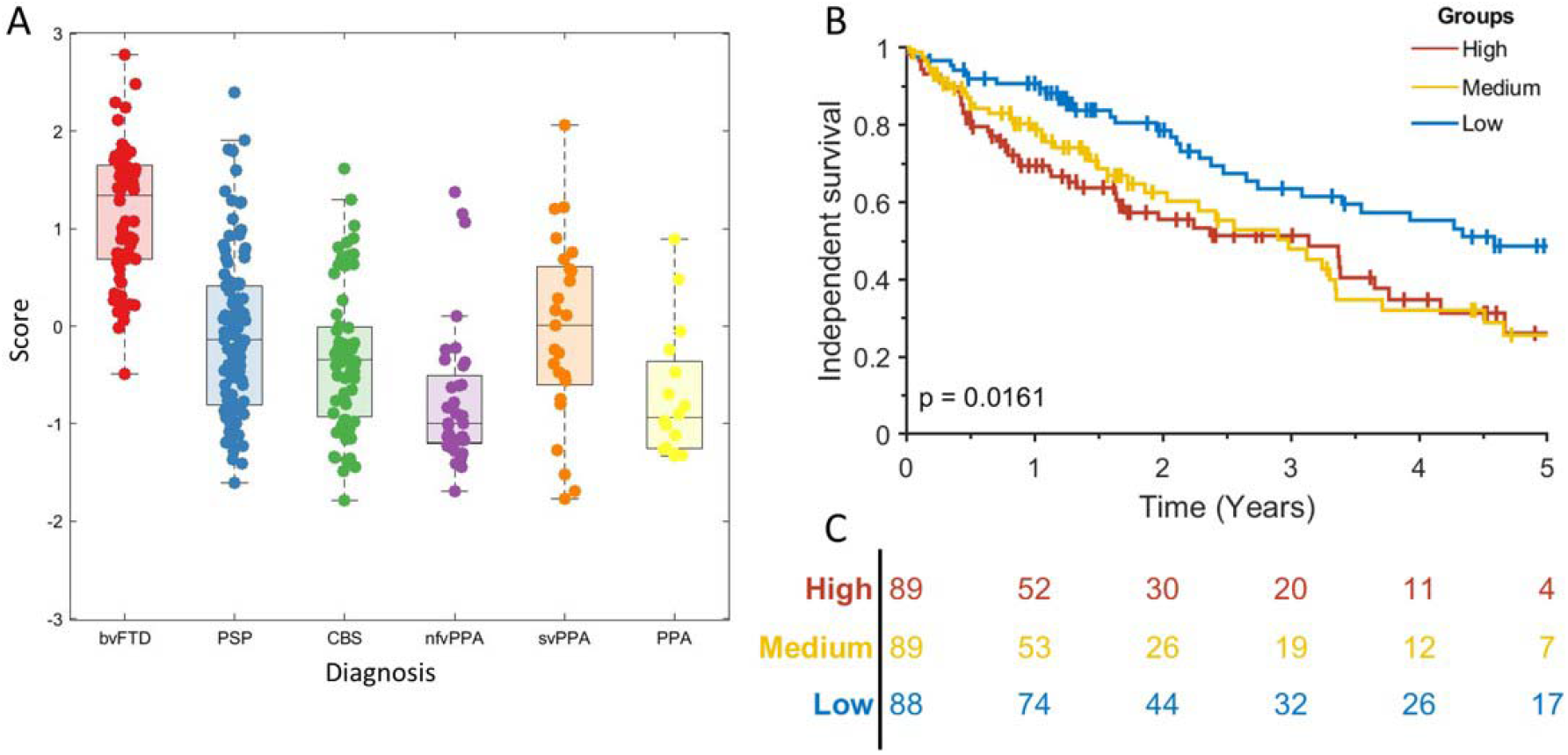
Independent survival (time to care home admission) in frontotemporal lobar degeneration syndromes. 3A: Scatterboxplot of each participant’s score on syndrome dimension one. 3B: Kaplan Meier survival curve for high, medium and low scoring tertiles for syndrome dimension one. The p value is from a log rank test of the null hypothesis of no difference in survival between all groups. Vertical lines show censored data. 3C: At risk table for the data shown in 3B.

Next, we tested which syndrome dimensions predicted care home admission at two years with age, gender and disease group as covariates. Eighty-nine patients were excluded from this analysis due to incomplete follow up. Syndrome dimension 1, reflecting behavioural impairment, was associated with care home admission (OR 2.46, p<0.001) (Table 4). This remained a significant predictor of care home admission even after bvFTD, the subgroup with highest scores, was removed (OR 3.20 p=0.03). Independent survival (time from clinical assessment to care home admission or death) differed between participants in high, medium and low severity tertiles for syndrome dimension 1 severity score (log rank p=0.007) (Figure 4). This result persisted after removing the bvFTD group (logrank p<0.001).

**Table 4:** Logistic regression of predictors of care admission by 2 years from clinical assessment.

|  | Odds ratio | Coefficient | *t* value | p value |
| --- | --- | --- | --- | --- |
| Constant | 0.10 | -2.26 | -1.17 | 0.24 |
| Age | 1.02 | 0.02 | 0.79 | 0.43 |
| Gender | 0.78 | -0.25 | -0.60 | 0.55 |
| Diagnosis 1 | 0.79 | -0.23 | -0.20 | 0.84 |
| Diagnosis 2 | 2.89 | 1.06 | 1.40 | 0.16 |
| Diagnosis 3 | 0.50 | -0.70 | -0.72 | 0.47 |
| Diagnosis 4 | 0.13 | -2.02 | -1.54 | 0.12 |
| Diagnosis 5 | 0.28 | -1.28 | -1.07 | 0.28 |
| **Syndrome dimension 1** | **2.46** | **0.90** | **3.11** | **<0.01** |
| Syndrome dimension 2 | 1.42 | 0.35 | 1.60 | 0.11 |
| Syndrome dimension 3 | 1.13 | 0.12 | 0.28 | 0.78 |
| Syndrome dimension 4 | 0.99 | -0.01 | -0.03 | 0.98 |
| Syndrome dimension 5 | 1.08 | 0.08 | 0.36 | 0.72 |
| Syndrome dimension 6 | 0.77 | -0.26 | -1.20 | 0.23 |

## Discussion

Clinician- and carer-rated behavioural disturbance is associated with shorter functionally independent survival, and the presence of motor features (including parkinsonism, postural instability, supranuclear gaze palsy, dystonia and apraxia) is associated with reduced absolute survival. These associations are found across the spectrum of common syndromes associated with FTLD, even when groups classically associated with these clinical features (bvFTD, PSP and CBS respectively) are excluded. A transdiagnostic approach that captures the clinical overlap and mixed phenotype adds clinically relevant information for prognostication to that available from the diagnostic group label.

Behavioural impairment, represented here by syndrome dimension 1, was associated with a greater risk of care home admission. This complements previous findings in bvFTD [5], Parkinson’s and Alzheimer’s disease [27,28]. Syndrome dimension 1 reflects behavioural impairments including apathy, impulsivity, socially inappropriate behaviour and hyperorality. More detailed neuropsychological tests and measures of carer burden could fractionate behavioural impairment to more closely determine which behavioural impairments have the greater effect on prognosis [8,29,30]. These results show correlation and not causation, and we lack data on the reasons given for care home admission. Behavioural impairments in frontotemporal dementia and PSP increase carer burden [31,32] and there are no proven effective pharmacological treatments [33,34]. Patients with more severe behavioural impairments may require continuous supervision which becomes difficult for spouses or families to sustain at home. We suggest that treating behavioural disturbance may delay the need for care home admission, with benefits to individual health and health-economics. Potential strategies include restoration of neurotransmitter deficits associated with behavioural change [35][36–38] or motor impairment [39,40].

The relationship between cognitive impairment and prognosis is complex. Some studies show a clear association [9], but others do not [8,41]. This discrepancy may be due to the indirect contribution of behavioural and motor impairments to performance on ‘cognitive’ tests. For example, speech or constructional deficits in nfvPPA or CBS respectively may impair performance tasks that require spoken, written or drawn response. However, the separation of cognitive and motor deficits across the six dimensions argues against such a simple interference effect.

The clinical phenotype reflected by syndrome dimensions 3 and 4 are classically associated with PSP Richardson’s syndrome and CBS respectively, but in our cohort was also expressed to a subtler degree by many other patients except for those with svPPA (Figure 2C&D). PSP-RS typically has a worse prognosis then bvFTD (unless there is coexistent MND) and PPA [3,42] while FTLD-tau has a worse prognosis than FTLD-TDP43 if clinical MND cases are excluded [43]. With disease progression many patients with nfvPPA develop the phenotype of PSP or CBS, which is an adverse prognostic sign [44–47]. In keeping with these observations, previous survival analyses of frontotemporal dementia (bvFTD and PPA) have shown that reduced letter fluency, motor cortex atrophy and brainstem hypoperfusion were associated with reduced survival [5,48,49]. Our results go beyond these findings, suggesting that development of motor impairments, irrespective of diagnostic group, is an adverse prognostic sign. However, the correlation between syndrome dimensions 3 and 4 and mortality does not prove causation. It is unclear if that these syndrome dimensions are indicative of a more aggressive disease or increased risk of complications, such as aspiration pneumonia due to dysphagia, sarcopenia and other aspects of frailty [50,51]. These complications could in turn increase mortality.

Our study has several limitations. We only included basic covariates in our survival analysis (age, gender and main diagnostic group). Medical and psychiatric comorbidities, marital status, social class, ethnicity and financial status are also known to influence rates of care home admission and death [28,52] and may explain some of the variance in prognosis. We attempted to recruit all patients with a designated syndrome associated with FTLD in our catchment area. Most referrals came from secondary care, so survival rates could be overestimated if patients with rapidly progressively disease died before they came to review. However average survival in our FTLD cohort was similar to those published previously [3]. We did not distinguish between residential or nursing care from basic demographic information. This was not differentiated in our demographic data because many institutions provide both levels of care at the same site. We also highlight that admission to a residential or nursing home is not a sign of inadequate home care and not inevitably associated with reduced quality of life. Patients can benefit from skilled holistic care provided in these institutions [53]. However, at a group level care home admission is a measure of reduced independence, and a potential study end point in trials.

In summary, functionally independent and absolute survival in syndromes associated with frontotemporal lobar degeneration are predicted by a subset of clinical features, over and above the diagnostic label. Given these findings, and the overlapping clinical [54], structural [7,55,56], functional [57,58], neuropathological [59,60] and neurochemical [35] features in these syndromes, we recommend a transdiagnostic approach to develop better treatment strategies. Effective treatments for behavioural and motor features could improve functionally independent survival and might reduce absolute mortality.

## Data Availability

Anonymised data are available on reasonable request for academic purposes.

## Acknowledgements

We would like to thank the patients and their families and carers and all the staff at the Cambridge Centre for Frontotemporal Dementia and Related Disorders, University of Cambridge.

